# COVID-19 vaccine effectiveness against hospitalizations and ICU admissions in the Netherlands, April- August 2021

**DOI:** 10.1101/2021.09.15.21263613

**Authors:** Brechje de Gier, Marjolein Kooijman, Jeanet Kemmeren, Nicolette de Keizer, Dave Dongelmans, Senna C.J.L. van Iersel, Jan van de Kassteele, Stijn P. Andeweg, the RIVM COVID-19 epidemiology and surveillance team, Hester E. de Melker, Susan J. M. Hahné, Mirjam J. Knol, Susan van den Hof

## Abstract

The objective of this study was to estimate vaccine effectiveness (VE) against COVID-19 hospitalization and ICU admission, per period according to dominating SARS-CoV-2 variant (Alpha and Delta), per vaccine and per time since vaccination. To this end, data from the national COVID-19 vaccination register was added to the national register of COVID-19 hospitalizations. For the study period 4 April – 29 August 2021, 15,571 hospitalized people with COVID-19 were included in the analysis, of whom 887 (5.7%) were fully vaccinated. Incidence rates of hospitalizations and ICU admissions per age group and vaccination status were calculated, and VE was estimated as 1-incidence rate ratio, adjusted for calendar date and age group in a negative binomial regression model. VE against hospitalization for full vaccination was 94% (95%CI 93-95%) in the Alpha period and 95% (95%CI 94-95%) in the Delta period. The VE for full vaccination against ICU admission was 93% (95%CI 87-96%) in the Alpha period and 97% (95%CI 97-98%) in the Delta period. VE was high in all age groups and did not show waning with time since vaccination up to 20 weeks after full vaccination.

## Introduction

Up until 29 August 2021, an estimated 82.8% of all Dutch inhabitants age 12 or older had received a first dose and 73.3% had completed their vaccination schedule (1, 2). The four COVID-19 vaccines used in the Netherlands are Comirnaty (BNT162b2; BioNTech/Pfizer, Mainz, Germany/New York, United States (US)), Spikevax (mRNA-1273, Moderna, Cambridge, US), Vaxzevria (ChAdOx1-S; AstraZeneca, Cambridge, United Kingdom) and Janssen (Ad26.COV2-S (recombinant), Janssen-Cilag International NV, Beerse, Belgium). The median interval between doses is 5 weeks for Comirnaty and Spikevax, and 11 weeks for Vaxzevria. Earlier reports have shown high effectiveness against SARS-CoV-2 infection and transmission in the Netherlands, but these studies were from a period where the Alpha variant (B.1.1.7) dominated (3, 4). Since July 2021, the Delta variant (B.1.617.2) is dominant in the Netherlands (5). Reports have shown a reduced effectiveness against SARS-CoV-2 Delta infection (6-8). However, a more important question is whether vaccine effectiveness against severe COVID-19 resulting in hospitalization or even ICU admission has diminished since the emergence of the Delta variant. Also, monitoring the possible waning of vaccine-induced immunity against severe COVID-19 is vital to estimate the burden on health care during autumn and winter 2021-2022, and to appraise the need for additional interventions such as booster doses.

In this study, we estimate incidence rates of COVID-19 hospitalizations and ICU admissions by vaccination status and age group to calculate vaccine effectiveness against COVID-19 hospitalizations and ICU admissions by age group, vaccine type, time since vaccination, and prevailing variant (Alpha or Delta).

## Methods

The number of hospitalizations and ICU admissions per vaccination status per day (the numerator) is based on a nationwide registry of COVID-19 hospitalizations (NICE)(9), linked to vaccination data from the national COVID-19 vaccination registry (CIMS), using the social services number as key to link the data. On request of the government all hospitalized persons with a positive SARS-CoV-2 test or CT-confirmed COVID-19 are registered in the NICE COVID-19 registry, a minimal dataset is made available to the RIVM for public health surveillance. For each person, the first admission date since June 2020 was included. In the Netherlands, every vaccinated person is asked for informed consent to have their vaccination data centrally registered in CIMS (COVID vaccination information and monitoring system). Therefore CIMS does not provide a complete registry of all administered COVID-19 vaccines. For people who are not registered as vaccinated in CIMS, we cannot be certain whether they are unvaccinated or whether they did not give informed consent. Of people vaccinated by the municipal health services (MHS), we know that 7.3% did not consent to registration in CIMS.

The number of persons per vaccination status in the population (the denominator) is based on a combination of vaccination registries: CoronIT, containing anonymized data on all vaccinations administered by the MHS (84% of all vaccinations in the Netherlands), supplemented with CIMS data for vaccinations by others, such as general practitioners and hospitals. The number of partly and fully vaccinated persons per age group and day was subtracted from the population size to calculate the size of the unvaccinated population.

Vaccination status was defined as ‘partly vaccinated’ 14 days after a first vaccine dose and becomes ‘fully vaccinated’ 14 days after a second dose or 28 days after the Janssen 1-dose schedule. People who received no vaccine or one dose less than 14 ago are categorized as unvaccinated. For hospitalized patients, the vaccination status on the date of disease onset was determined. The NICE data does not contain disease onset dates. Therefore, the median number of days between disease onset and admission date per age group (ranging between 3-7 days) was retrieved from the national notification database Osiris-AIZ and applied to admission dates in NICE to estimate disease onset dates. The dataset included hospitalizations with admission dates until September 5, for this analysis we included hospitalizations with an estimated date of disease onset up to August 29, 2021.

Incidence rates per 10.000 person-days were calculated by dividing the number of hospitalizations and ICU admission by the population size, per vaccination status. Incidence rate ratio’s (IRR) and 95% confidence intervals (95%CI) were estimated with a negative binomial regression model with log-link function, to allow for overdispersion in the numbers. The number of hospitalizations or ICU admissions was the dependent variable, vaccination status, 5-year age group and calendar dates were independent variables. Calendar date was included as penalized spline to model the non-linear effect of time on incidence rates. The logarithm of population size was included as offset. Vaccine effectiveness (VE) was calculated as 1-IRR x 100%.

All VE estimates were stratified in two periods, when a variant of concern (VOC) was clearly dominant in the Netherlands. The Alpha period is 4 April 2021– 29 May 2021, when over 95% of weekly sequenced SARS-CoV-2 isolates pertained the Alpha variant. The Delta period is 4 July 2021 -29 August 2021, which is the end of the study period, with 99.9% of all sequenced isolates pertaining the Delta variant (5).

## Results

A total of 16,162 persons were hospitalized with COVID-19 with an estimated disease onset date between 4 April and 29 August 2021. Of these, 591 (3.7%) were excluded because they could not be linked to a valid social services number. Of 15,571 included patients, 887 (5.7%) were fully vaccinated on the date of disease onset 1,111 (7.1%) were partly vaccinated and 13,574 (87.2%) were unvaccinated. Of the 13,574 unvaccinated patients, 7,406 were found in the CIMS registry with a vaccination date after hospitalization. Of the other 6,168 hospitalized patients, no vaccination records were found in CIMS.

Figure 1a shows that most hospitalized persons were unvaccinated. From July onward, the number of vaccinated patients approaches the number of unvaccinated patients in the age group 70 years and older. However, Figure 1b shows that the incidence rate of hospitalizations per 100.000 person-days is far lower among vaccinated than among unvaccinated persons. The similar numbers in absolute terms in the age group 70 years and older are due to the high vaccination coverage in this group. With increasing vaccination coverage, the incidence rate in the fully vaccinated population approaches the overall incidence rate in the population (Figure 1b). The same trends are visible for ICU admissions (Figure 2a and 2b), although the absolute number of ICU admissions in vaccinated persons is still well below the number of ICU admissions in unvaccinated persons.

**Figure 1.**
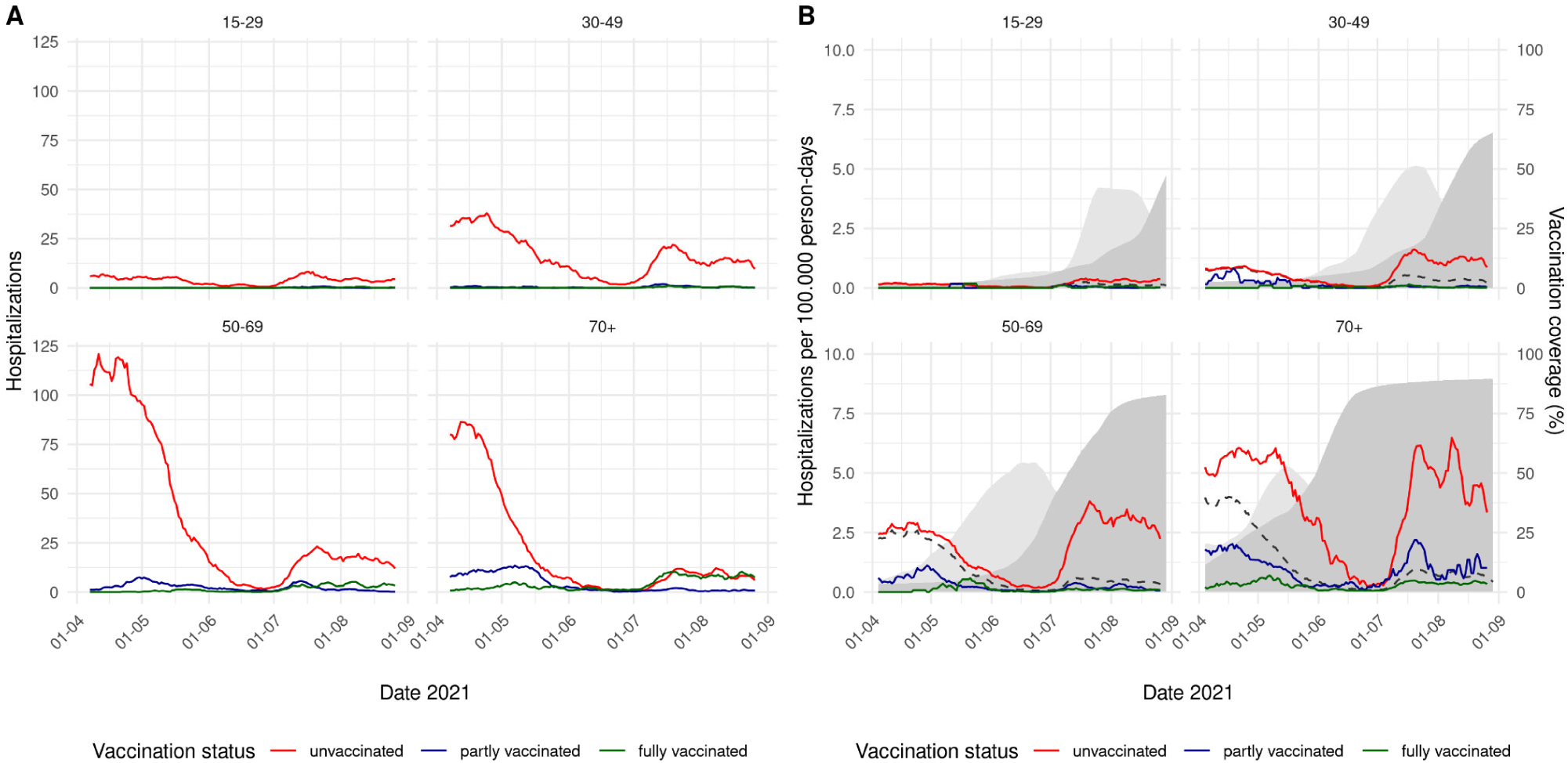
A. 7-day moving average of the number of COVID-19 hospitalizations by vaccination status and age group, 4 April – 29 August 2021. B. 7-day moving average of the incidence rate of COVID-19 hospitalizations per 100.000 person-days by vaccination status and age group, 4 April – 29 August 2021. The dashed line shows the overall incidence in the age-specific population. The light grey area shows the percentage of partly vaccinated persons in the population, the dark grey area the percentage of fully vaccinated persons.

**Figure 2.**
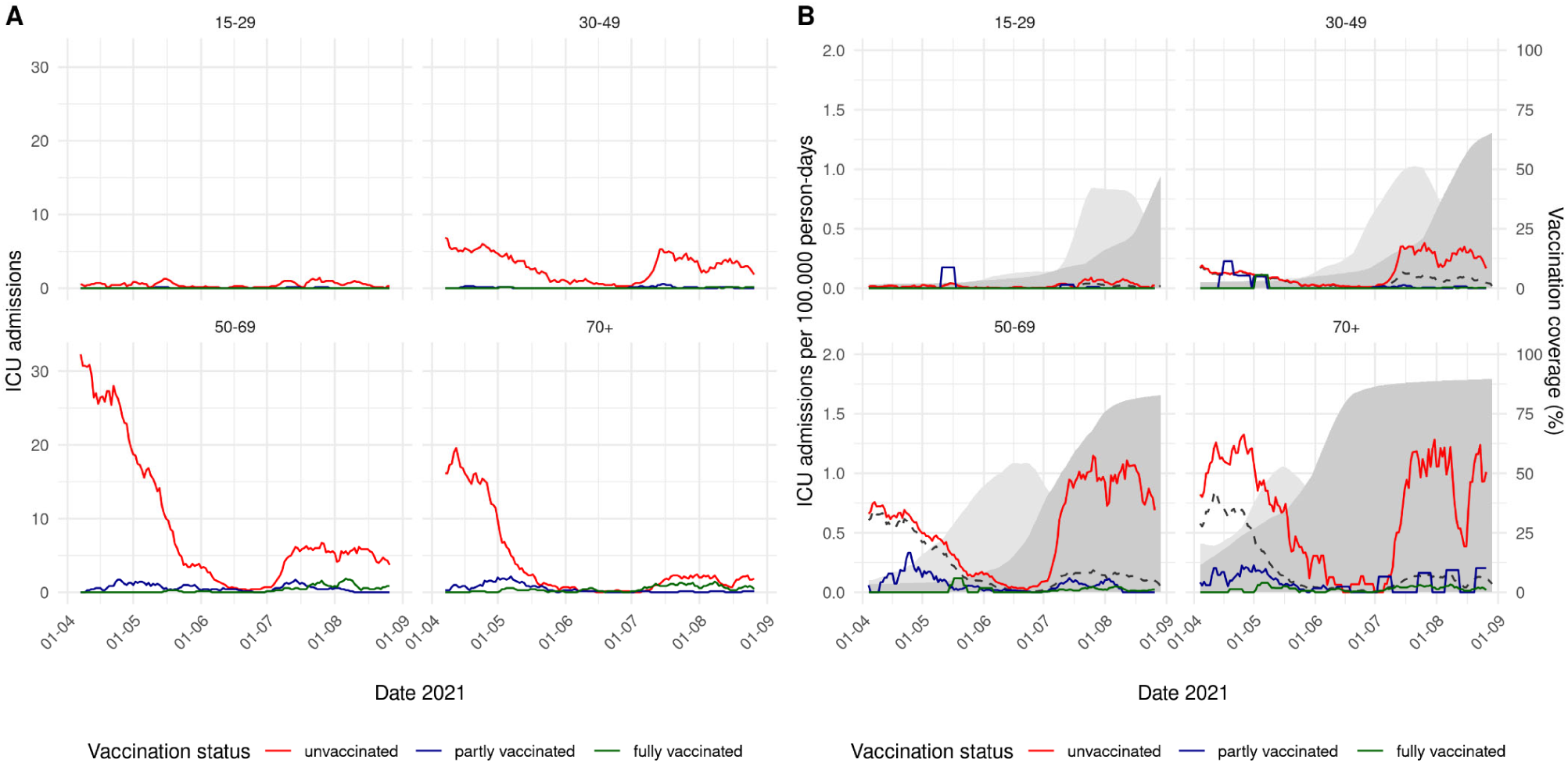
A. 7-day moving average of the number of COVID-19 ICU admissions by vaccination status and age group, 4 April – 29 August 2021. B. 7-day moving average of the incidence rate of COVID-19 ICU admissions per 100.000 person-days by vaccination status and age group, 4 April – 29 August 2021. The dashed line shows the overall incidence in the age-specific population. The light grey area shows the percentage of partly vaccinated persons in the population, the dark grey area the percentage of fully vaccinated persons.

Table 1 shows the estimated VE against hospitalizations and ICU admissions by age group, for the Alpha and Delta periods. The overall VE for full vaccination against hospitalization was 94% (95%CI 93-95%) in the Alpha period and 95% (95%CI 94-95%) in the Delta period. The overall VE for full vaccination against ICU admission was 93% (95%CI 87-96%) in the Alpha period and 97% (95%CI 97-98%) in the Delta period. High VEs for full vaccination were found for all age groups in both periods.

**Table 1.**
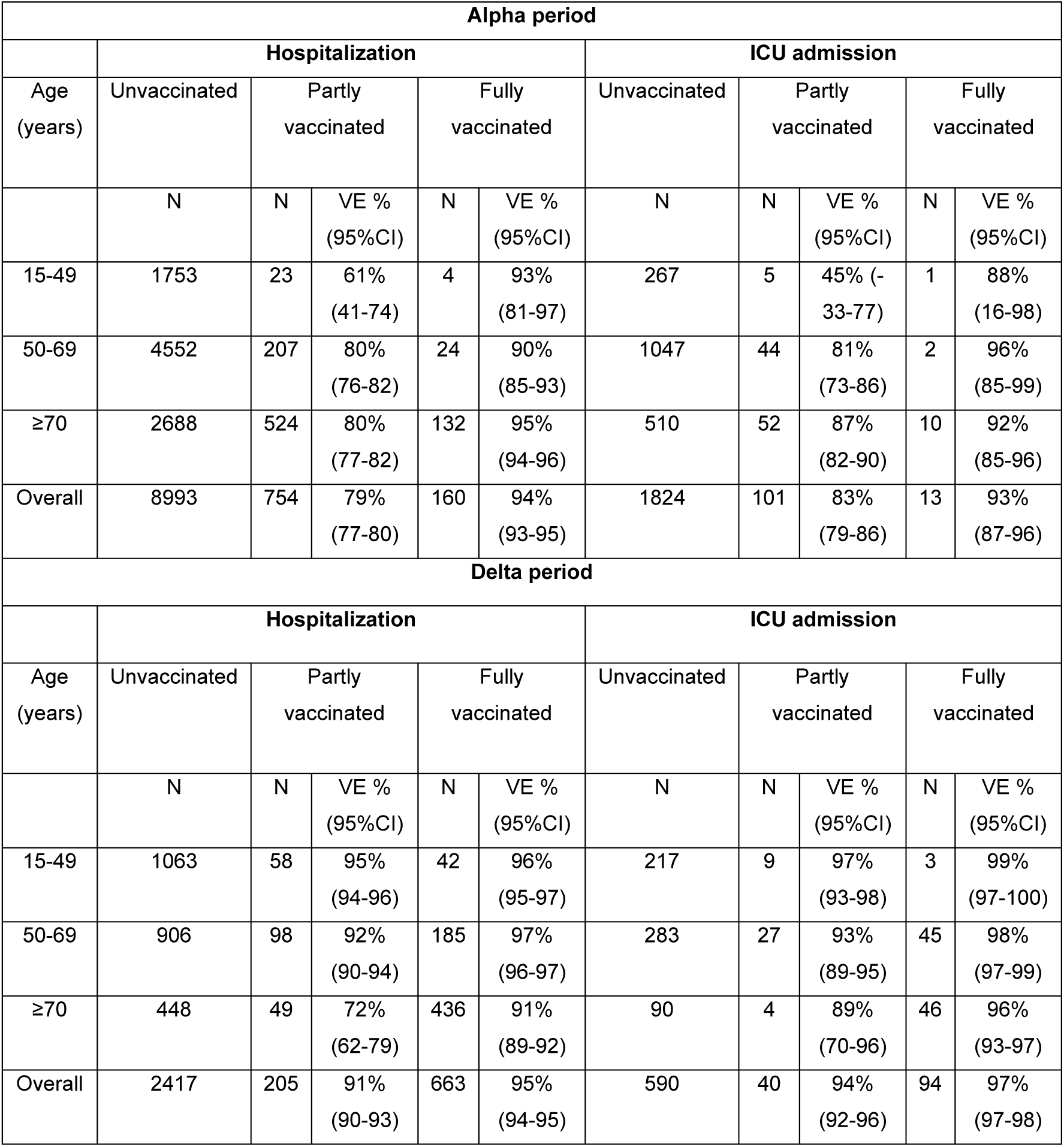
Vaccine effectiveness (VE) and 95% confidence intervals (CI) against hospitalization and ICU admission for the Alpha and Delta periods, overall and by age group. All VE estimates are adjusted for calendar date. The overall VE is also adjusted for age group.

| Alpha period |  |  |  |  |  |  |  |  |  |  |
| --- | --- | --- | --- | --- | --- | --- | --- | --- | --- | --- |
|  | Hospitalization |  |  |  |  | ICU admission |  |  |  |  |
| Age (years) | Unvaccinated | Partly vaccinated |  | Fully vaccinated |  | Unvaccinated | Partly vaccinated |  | Fully vaccinated |  |
|  | N | N | VE % (95%CI) | N | VE % (95%CI) | N | N | VE % (95%CI) | N | VE % (95%CI) |
| 15-49 | 1753 | 23 | 61% (41-74) | 4 | 93% (81-97) | 267 | 5 | 45% (-33-77) | 1 | 88% (16-98) |
| 50-69 | 4552 | 207 | 80% (76-82) | 24 | 90% (85-93) | 1047 | 44 | 81% (73-86) | 2 | 96% (85-99) |
| ≥70 | 2688 | 524 | 80% (77-82) | 132 | 95% (94-96) | 510 | 52 | 87% (82-90) | 10 | 92% (85-96) |
| Overall | 8993 | 754 | 79% (77-80) | 160 | 94% (93-95) | 1824 | 101 | 83% (79-86) | 13 | 93% (87-96) |
| Delta period |  |  |  |  |  |  |  |  |  |  |
|  | Hospitalization |  |  |  |  | ICU admission |  |  |  |  |
| Age (years) | Unvaccinated | Partly vaccinated |  | Fully vaccinated |  | Unvaccinated | Partly vaccinated |  | Fully vaccinated |  |
|  | N | N | VE % (95%CI) | N | VE % (95%CI) | N | N | VE % (95%CI) | N | VE % (95%CI) |
| 15-49 | 1063 | 58 | 95% (94-96) | 42 | 96% (95-97) | 217 | 9 | 97% (93-98) | 3 | 99% (97-100) |
| 50-69 | 906 | 98 | 92% (90-94) | 185 | 97% (96-97) | 283 | 27 | 93% (89-95) | 45 | 98% (97-99) |
| ≥70 | 448 | 49 | 72% (62-79) | 436 | 91% (89-92) | 90 | 4 | 89% (70-96) | 46 | 96% (93-97) |
| Overall | 2417 | 205 | 91% (90-93) | 663 | 95% (94-95) | 590 | 40 | 94% (92-96) | 94 | 97% (97-98) |

Table 2 shows the estimated VE per age group per vaccine, for the Delta period. Because of small numbers of fully vaccinated hospitalizations during the Alpha period, we do not present VE by vaccine type for this period. High VE are found for all four vaccines, with the exception of Spikevax in the oldest age group. Please note that in the Netherlands, persons with a medical condition putting them at high risk of severe COVID-19 were specifically given the Spikevax vaccine. The Janssen vaccine is not allocated to elderly in the Netherlands, therefore no VE for ≥70 was estimated. Table 3 shows the VE by time since the ‘fully vaccinated’ state was reached, in 5-week periods. No indication for VE waning is observed up to 20 weeks after full vaccination.

**Table 2.** Vaccine effectiveness (VE) and 95% confidence interval (CI) against hospitalization and ICU admissions, during the Delta period (4 July – 12 August 2021), per age group, per vaccine. All VE estimates are adjusted for calendar date and 5-year age group.

|  | Delta period |  |  |  |  |  |  |  |
| --- | --- | --- | --- | --- | --- | --- | --- | --- |
|  | Hospitalization |  |  |  | ICU admission |  |  |  |
|  | Partly vaccinated |  | Fully vaccinated |  | Partly vaccinated |  | Fully vaccinated |  |
| Age (years) | N | VE % (95%CI) | N | VE % (95%CI) | N | VE % (95%CI) | N | VE % (95%CI) |
| **Comirnaty® (BNT162b2)** |  |  |  |  |  |  |  |  |
| 15-49 | 50 | 95% (93-96) | 7 | 99% (98-100) | 8 | 96% (93-98) | 0 | 100% (--) |
| 50-69 | 60 | 92% (90-94) | 52 | 99% (98-99) | 18 | 93% (88-95) | 7 | 100% (99-100) |
| $\geq 70$ | 42 | 71% (60-79) | 391 | 92% (90-93) | 3 | 91% (70-97) | 36 | 97% (95-98) |
| Overall | 152 | 91% (90-93) | 450 | 96% (95-96) | 29 | 94% (91-96) | 43 | 99% (98-99) |
| **Spikevax® (mRNA-1273)** |  |  |  |  |  |  |  |  |
| 15-49 | 4 | 97% (92-99) | 22 | 88% (82-92) | 0 | 100% (--) | 1 | 98% (85-100) |
| 50-69 | 4 | 96% (90-99) | 43 | 89% (85-92) | 2 | 94% (75-98) | 14 | 89% (80-93) |
| $\geq 70$ | 1 | 75% (-79-96) | 29 | 64% (47-76) | 0 | 100% (--) | 10 | 34% (-29-66) |
| Overall | 9 | 96% (92-98) | 94 | 84% (80-87) | 2 | 97% (87-99) | 25 | 86% (79-90) |
| **Vaxzevria® (ChAdOx1-S)** |  |  |  |  |  |  |  |  |
| 15-49 | 1 | 88% (18-98) | 4 | 95% (87-98) | 0 | 100% (--) | 1 | 95% (64-99) |
| 50-69 | 32 | 90% (86-93) | 67 | 96% (95-97) | 7 | 93% (86-97) | 19 | 96% (94-98) |
| $\geq 70$ | 6 | 78% (50-90) | 16 | 78% (63-86) | 1 | 70% (-114-96) | 0 | 100% (--) |
| Overall | 39 | 88% (83-91) | 87 | 94% (92-95) | 8 | 92% (84-96) | 20 | 96% (94-98) |
| **Janssen® (Ad26.COV2-S)** |  |  |  |  |  |  |  |  |
| 15-49 | 3 | 88% (62-96) | 9 | 93% (86-96) | 1 | 77% (-62-97) | 1 | 96% (73-99) |
| 50-69 | 2 | 67% (-33-92) | 23 | 92% (89-95) | 0 | 100% (--) | 5 | 94% (86-98) |
| Overall | 5 | 82% (56-93) | 32 | 91% (88-94) | 1 | 83% (-22-98) | 6 | 94% (88-98) |

**Table 3.** Vaccine effectiveness (VE) and 95% confidence interval against hospitalization and ICU admissions in the Delta period, per age group and time since full vaccination (week 0 starts 14 days after a second dose of a two-dose schedule or 28 days after Janssen vaccination). All VE estimates are adjusted for calendar date and 5-year age group.

| Delta period |  |  |  |  |  |
| --- | --- | --- | --- | --- | --- |
| Age (years) | Time interval since full vaccination | Hospitalization |  | ICU admission |  |
|  |  | N | VE % (95%CI) | N | VE % (95%CI) |
| 15-49 | Unvaccinated | 1063 |  | 217 |  |
|  | 0-4 weeks | 12 | 99% (97-99) | 0 | 100% (--) |
|  | 5-9 weeks | 15 | 93% (88-96) | 1 | 98% (85-100) |
|  | 10-14 weeks | 12 | 75% (56-86) | 2 | 82% (29-96) |
|  | 15-19 weeks | 1 | 97% (76-100) | 0 | 100% (--) |
|  | 20 or more weeks | 2 | 97% (87-99) | 0 | 100% (--) |
| 50-69 | Unvaccinated | 906 |  | 283 |  |
|  | 0-4 weeks | 68 | 98% (97-98) | 13 | 99% (98-99) |
|  | 5-9 weeks | 70 | 97% (96-98) | 19 | 98% (97-99) |
|  | 10-14 weeks | 33 | 90% (85-93) | 9 | 93% (85-96) |
|  | 15-19 weeks | 9 | 92% (84-96) | 4 | 89% (70-96) |
|  | 20 or more weeks | 4 | 98% (94-99) | 0 | 100% (--) |
| ≥70 | Unvaccinated | 448 |  | 90 |  |
|  | 0-4 weeks | 26 | 90% (85-93) | 1 | 99% (93-100) |
|  | 5-9 weeks | 115 | 92% (90-93) | 26 | 95% (92-97) |
|  | 10-14 weeks | 144 | 90% (88-92) | 13 | 96% (93-98) |
|  | 15-19 weeks | 117 | 91% (88-92) | 3 | 97% (89-99) |
|  | 20 or more weeks | 30 | 91% (87-94) | 2 | 90% (57-98) |

## Discussion

This study using nationwide surveillance data found 95% and 97% effectiveness of the COVID-19 vaccines used in the Netherlands against hospitalization and ICU admission, respectively. This high VE persisted into the Delta period and over 20 weeks after full vaccination. Our findings are in line with previous studies showing that while VE against SARS-CoV-2 infection is lower with Delta compared to Alpha variant, the VE against severe disease remains very high (10). A study from Israel did indicate possible waning over time of VE against severe COVID-19 over time (11). In our data, adjusting for 5-year age groups, we did not observe such waning up to 20-plus weeks after full vaccination. Close monitoring of VE against severe disease by time since vaccination is important to timely detect a possible need for additional intervention such as booster doses.

Linkage of the NICE COVID-19 hospitalization registry with the CIMS vaccination registry allowed for a nationwide analysis of VE against hospitalization, per vaccine and per time since vaccination. However, this dataset has three important limitations. Firstly, the NICE COVID-19 dataset does not contain information on comorbidities. When interpreting our results, it is important to consider that groups with increased risk of severe COVID-19 were prioritized for vaccination in early 2021. The relatively low VE of Spikevax (Moderna) might be related to the use of specifically this vaccine for patient groups at medically high risk for severe COVID-19, such as immunocompromised patients. In this group, the chance of vaccine breakthrough infection requiring hospitalization is relatively high (12-14). Other studies have not found a lower VE against severe COVID-19 for the Spikevax vaccine in the general population. Puranik et al. found a Spikevax VE against hospitalization of 81% in the US in July 2021, and a Canadian study found a single-dose Spikevax VE against hospitalization with Delta of 96% (6, 15).

Secondly, the NICE COVID-19 registry includes all hospitalized patients with a positive SARS-CoV-2 test, also when the indication for hospitalization was not COVID-19, when patients test positive by routine screening at admission. This might bias our VE estimates if vaccination impacts the likelihood of being admitted with SARS-CoV-2 rather than because of SARS-CoV-2. Such misclassification of COVID-19 hospitalizations could be more likely for vaccinated patients as their risk of severe COVID-19 is greatly reduced resulting in underestimation of the VE. On the other hand unvaccinated patients have a higher risk of being infected in general, which could lead to overestimation of the VE. A third limitation is the incompleteness of the CIMS registry, due to the required informed consent of vaccinees. Because of this incompleteness, some patients without vaccination records in CIMS might be misclassified as unvaccinated. This could lead to an overestimation of VE. We explored the possible influence of this misclassification on our estimates with a simple sensitivity analysis, assuming misclassification of 7.3% of all truly vaccinated cases as unvaccinated. This reduced the VE estimates by only 1 -2%.

A limitation of the method used is that previously infected persons who have developed natural immunity are not removed from the person-time at risk. A recent study showed that 20-25% of Dutch blood donors has acquired immunity through infection (16). Further, confounding by unmeasured factors associated with odds of both vaccination and hospitalization may have influenced our estimates. For example, some reports have suggested a lower vaccination coverage in Dutch inhabitants with a migration background and also a high proportion of hospitalized patients from this sub-population (17, 18). Lastly, our estimates per vaccine and per time since vaccination are sometimes imprecise due to low numbers.

In conclusion, the COVID-19 VE against hospitalization and ICU admissions are very high in the Netherlands, also in the period when the Delta variant dominates and up to 20 weeks after full vaccination. The observed lower protection of persons vaccinated with the Spikevax vaccine is likely a result of the allocation of this vaccine to the population at medical high risk of severe COVID-19 and of vaccine breakthrough infection. Additional interventions might be needed to protect this vulnerable group against severe COVID-19. Among the unvaccinated population, the incidence of COVID-19 hospitalizations and ICU admissions is high, which, combined with the size of this population, could lead to a significant strain on health care in the coming months. A further increase in vaccination coverage is key to reduce the burden on health care.

## Data Availability

The primary data is not publicly available. Aggregated tables of these data are made available weekly at the website of the RIVM.

## Acknowledgments

*The data used in this study are collected, maintained and disclosed by the MHS (GGDen), GGDGHOR Nederland, RIVM Dienst Vaccinvoorziening en Preventie, all Dutch hospitals and the NICE regsitration*.

## Members of the RIVM COVID-19 surveillance and epidemiology team

Agnetha Hofhuis, Anne Teirlinck, Alies van Lier, Bronke Boudewijns, Carolien Verstraten, Gert Broekhaar, Guido Willekens, Irene Veldhuijzen, Jan Polman, Jan van de Kassteele, Jeroen Alblas, Janneke van Heereveld, Janneke Heijne, Kirsten Bulsink, Lieke Wielders, Liselotte van Asten, Liz Jenniskens, Loes Soetens, Maarten Mulder, Maarten Schipper, Marit de Lange, Naomi Smorenburg, Nienke Neppelenbroek, Patrick van den Berg, Priscila de Oliveira Bressane Lima, Rolina van Gaalen, Sara Wijburg, Shahabeh Abbas Zadeh Siméon de Bruijn, Senna van Iersel, Stijn Andeweg, Sjoerd Wierenga, Susan Lanooij, Sylvia Keijser, Tara Smit, Don Klinkenberg, Jantien Backer, Pieter de Boer, Scott McDonald, Amber Maxwell, Annabel Niessen, Brechje de Gier, Danytza Berry, Daphne van Wees, Dimphey van Meijeren, Eric R.A. Vos, Frederika Dijkstra, Jeanet Kemmeren, Kylie Ainslie, Marit Middeldorp, Marjolein Kooijman, Mirjam Knol, Timor Faber, Albert Jan van Hoek, Eveline Geubbels, Birgit van Benthem, Hester de Melker, Jacco Wallinga, Rianne van Gageldonk-Lafeber, Susan Hahné, Susan van den Hof

